# Effects of a School-Based Physical Activity Intervention on Children with Intellectual Disability: A Cluster Randomised Trial

**DOI:** 10.1101/2024.12.22.24319526

**Authors:** Michael Noetel, Taren Sanders, Danielle Tracey, David R. Lubans, Viviene A Temple, Andrew Bennie, James Conigrave, Mark Babic, Bridget Booker, Rebecca Pagano, James Boyer, Chris Lonsdale

## Abstract

**Importance:** Young people living with disability have poorer health outcomes than their typically developing peers. They are less physically active and at increased risk of chronic disease. Teacher-led, whole-of-school physical activity interventions are promising levers for population-level change, but are seldom tested among children with disability.

**Objective:** To evaluate the effect of a blended teacher-professional learning program (online and in-person) on fundamental movement skills among children with disability.

**Design, Setting, and Participants:** In this cluster randomised clinical trial, we randomised 20 government-funded primary schools, including 238 consenting students between Grades 2-5. Ten schools received the blended teacher-professional learning intervention and 10 received the control intervention. The professional learning was designed to support teachers as they implemented a whole-of-school intervention designed to enhance fundamental movement skills and increase physical activity levels. Recruitment and baseline assessments occurred in 2020. Research assistants, blinded to treatment allocation, completed follow-up outcome assessments at 21 months.

**Interventions:** The school-based intervention was mostly online learning for teachers, followed by one lesson observation from a project mentor and one from a peer. Between one and three teachers also met quarterly with the mentor to plan whole-of-school strategies to promote activity.

**Main Outcomes and Measures:** Test of Gross Motor Development-3 test of fundamental movement skill competency. Secondary outcomes were self-concept, enjoyment, wellbeing, 300-yard run time, and accelerometer-measured physical activity.

**Results:** We found no significant group-by-time effects for the primary outcome (fundamental movement skill competency: b = 1.07 [95% CI -3.70, 5.84], p = .658) or any of the secondary outcomes. About half the teachers assigned to the training completed the modules.

**Conclusions and Relevance:** In this study, a school-based intervention did not improve children’s fundamental movement skill competency or any other outcomes. Results may be attenuated by the disruptive effects of the COVID-19 pandemic due to reduced teacher participation and delayed post-test measurement. Alternatively, low intensity teacher-professional learning interventions may not be enough to improve motor competence or physical activity among children with intellectual disability.

**Key Points:** *Question:* Does a blended-learning intervention for teachers improve fundamental movement skills among children with intellectual disability?

*Findings:* In this cluster randomised clinical trial of 20 schools and 238 children, a blended teacher-professional learning intervention did not improve fundamental movement skill competency at 21 months.

*Meaning:* Given similar programs increased outcomes among typically developing children, results of this randomised clinical trial suggest different models are needed to support children with disability.

---

Around 15% of the global population live with disability.^1^ People with disabilities often show great resilience and adaptability.^2^ They also face challenges, typically having poorer health outcomes than those without;^1^ for example, those with intellectual disabilities die 20 years earlier than their neurotypical peers.^3^ The World Health Organisation (WHO) attributes a large part of this mortality risk to higher rates of noncommunicable diseases among those with disability.^4^ It estimates a $10 return for every $1 spent on disability-inclusive prevention of non-communicable diseases.^4^

A potent lever for preventing non-communicable diseases is increasing physical activity, particularly among young people.^5^ According to the latest physical activity global report card, young people in most countries fail to meet physical activity guidelines, with a global average fail rate around 70%.^6^ The risk of being insufficiently active is higher for children and adolescents with disability.^7,8^ Physical activity also has more immediate benefits for children with disability, increasing physical and psychosocial health indicators.^8,9^ As a result, the WHO recommends the same amount of physical activity for children with and without disability.^5^

Schools are key settings for increasing physical activity.^10^ One billion children attend school each day, and they spend more time at school than anywhere except home.^10^ Whole-of-school physical activity interventions have been shown to increase health and physical activity among typically developing students.^10^ However, there is limited research on effective physical activity interventions for children with disability.^11,12^ A systematic review on children with physical disability found only seven studies, all of which were on cerebral palsy, and none of which were in schools.^12^ A review on children with intellectual disability found only five studies,^11^ only one of which was in schools.^13^ The one school-based intervention was found to increase health indicators among adolescents with intellectual disability.^14^ Our research aims to see if similar benefits can be achieved for children.

Our current study builds upon three projects for children without disability.^15–17^ These studies used whole-of-school interventions to increase cardiorespiratory fitness,^15,16^ physical activity,^15,17^ wellbeing,^17^ and movement skill competency among children.^15^ By using principles from the Consolidated Framework for Implementation Research,^18^ the intervention could be delivered for 26 USD/student.^16^ For the same reasons, many effects were maintained when scaling-up the intervention to 115 schools.^17^

In this project, we adapted the intervention to support *all* children, including those with disability. We focused on measuring effects among children with intellectual disability because it was the most common type of disability among school students (65% among boys, 54% among girls).^19^ We tested the hypothesis that this teacher professional learning intervention increased fundamental movement skill (FMS) competency among those children. These movement skills are core learning objectives of primary school physical activity,^20^ and a major component of physical literacy.^21^ These skills can be reliably assessed in children with intellectual disability.^22^ They predict physical activity, cardiorespiratory fitness, and weight status in typically developing children.^23,24^ They also predict many health and developmental outcomes in children with intellectual disabilities.^22^ As secondary outcomes, we also assessed whether the intervention increased physical activity, physical fitness, confidence, enjoyment, and wellbeing.

## Methods

### Trial Design and Participants

We received ethical approval from both the Australian Catholic University Human Research Ethics Committee (approval 2019-106H) and the NSW Department of Education (State Education Research and Partnerships number 2019289). We used a cluster-randomised trial, nesting students within schools. All government primary schools within 3 hours of our university were eligible, as long as the school had at least 10 students between Years 2 and 5 with intellectual disabilities. At consenting schools, all children with intellectual disabilities between those ages were eligible for data collection, with two exceptions. Our measures were not appropriate for students whose disability prevented them from running (e.g., wheelchair users) or those whose comorbid developmental disorders precluded them from responding to verbal questions (e.g., level 2 and 3 Autism Spectrum Disorder). We made one major change to the protocol following registration: we originally scheduled post-test data collection for 12 months after baseline. At this point in time, COVID-19 led the state government to prohibit data collection in schools for high-risk groups, including those with intellectual disabilities. Rather than abandoning data collection, we followed recommendations to maintain trial integrity, where possible.^25^ To allow restrictions to abate, we postponed our endpoint data-collection for three school terms (∼9 months), making our post-test 21 months after baseline. Given this period spanned two school years, we could not meaningfully nest students within teachers for our main analyses.

### Interventions

Full details of our intervention are set out in our protocol.^26^ We adapted the ‘internet-based Professional Learning to help teachers promote Activity in Youth’ (iPLAY)^16,17,27^ intervention for children with disability. The iPLAY program helps schools implement quality physical education (PE) and school sport, classroom energisers, physically active homework, active playgrounds, links to community physical activity, and parent engagement. We consulted with special education teachers and experts in intellectual disability to identify how iPLAY should best be adapted for this population. To better support students with disability, iPLAY for Inclusion (iPLAY4i) abbreviated some content that was likely less relevant to these schools (e.g., active homework, where many schools for this population had ‘no homework’ policies). Instead we added content on positive behaviour support^28^ and universal design for learning.^29^ For example, we trained teachers in the TREE framework where teachers introduce variations (to the Teaching style, Rules, Equipment or Environment)^30^ that allow all students to experience success during school sport and physical education sessions.

Classroom teachers delivered curricular components (e.g., quality PE and school sport, classroom energises) which were built around making classes SAAFE:^31^ Supportive, Active, Autonomous, Fair and Enjoyable. Up to three classroom teachers delivered the non-curricular components of the intervention (e.g., active playgrounds). These ‘iPLAY leaders’ also supported other teachers with implementation of curricular components. As per previous studies, ^27^ schools adopted these strategies with support from a two-hour face-to-face workshop, online resources, and visits from an ‘iPLAY mentor’—an experienced physical education teacher employed by the project team. Those visits provided classroom teachers with one lesson observation, and provided leaders with quarterly planning sessions for their non-curricular components.

### Outcomes

All outcomes were measured at baseline and at 21 months post-test.

### Primary Outcome: Test of Gross Motor Development-3 (TGMD-3)^32^

The TGMD-3 has been validated for children with developmental disorders, including intellectual disabilities.^33–35^ Due to time constraints, we chose the three skills for each subscale that explained the most variance in children with intellectual disabilities (run, gallop, and hop for locomotor skills; one-handed strike, dribble, and kick for object-control skills).^35,36^ Students were provided with a physical demonstration by a research assistant, a practice attempt, and a set of visual prompts shown to increase reliability and validity.^33^ Students completed each skill two times and were video-recorded. The outcome was coded by a research assistant who was blind to the treatment allocation. We coded 10% of the data in duplicate to assess inter-rater reliability. We pre-registered our decision to use the total score for the two attempts of these six skills.

### Secondary outcome measures

Measuring cardiorespiratory fitness in young people with intellectual disabilities is challenging. Most assessments fail to meet established reliability and validity standards.^37^ We measured cardiovascular fitness using the 300-yard run after pilot testing because: we wanted a short, simple measure, given our young sample; and the 300-yard run has been shown to be reliable in young people with intellectual disabilities.^37–39^ Children were asked to run or walk as fast as they can over 300 yards, timed by a research assistant.

We measured students’ physical activity over seven days using GENEActiv accelerometers, worn on the non-dominant wrist. Wrist-based accelerometry is valid and acceptable for children with intellectual disabilities.^40^ Data were reduced using the Euclidean norm minus one (ENMO) method to apply cut-points to the data, providing estimates of time in different intensities of activity (e.g., moderate vs. vigorous). We used school bell-times to examine physical activity within school, during breaks, after school, on weekends, and in total.

We asked students about their enjoyment of physical activity,^41^ their physical self-concept,^42,43^ and their satisfaction with life.^44^ We used individual administration, where researchers first ask students whether they agree with a statement, and then to what extent. We also included acquiescence checks to ensure that self-report responses were viable. These checks included the items “Do you make all your own clothes and shoes?” and “Where you live, did you choose who lives next door to you?” If participants responded yes to either of these questions they were excluded from the analyses with self-report outcomes. These methods lead to better validity among children with intellectual disability.^42^

### Sample Size

In typically developing children, a meta-analysis of interventions found a large pooled effect size on overall gross-motor competency.^20^ To estimate our effect size, we used the smallest pooled standardised mean difference reported in that meta-analysis (object control = .63). We used data from the Department of Education to estimate eligible students per school (∼20 students meeting criteria in Years 2-5; conservative 30% consent rate). We also conservatively estimated the sample size using post-test means, instead of a more statistically powerful ANCOVA. We used G*Power 3 to estimate the required sample size to achieve 80% power,^45^ then increased this value using a design effect^46^ (ICC = 0.08 from previous research).^15^ Given the above parameters, we needed 20 schools (10 intervention, 10 control) with 115 students total (5-6 students per school) to reach >80% power.

### Recruitment, Assignment, Randomisation, and Blinding

We matched schools on school type (regular vs. schools for specific purposes), Index of Community Socio-Educational Advantage (ICSEA), and location (urban vs. remote). Then, an experienced statistician who was not part of the research team used a computer-generated algorithm to randomise matched schools, 1:1 into treatment and control.^47^

Outcome assessors were blind to allocation. We were unable to blind teachers because they were aware of the training during recruitment for the trial. We did not tell students whether or not their school received the training, however we could not prevent staff from discussing the training with students. Nevertheless, we judged the likelihood of students’ awareness influencing results would be low.

### Statistical Analysis

We used R version 4.4.0 for our analyses.^48^ In line with intention-to-treat procedures, all available student data was retained and included in mixed model analyses, even in cases where students did not provide data for both timepoints.^49^ Multiple imputation (predictive mean matching with the *mice* package)^50^ to manage missing measurements (e.g., covariates) within a timepoint. We then used linear mixed models for intervention effects using *lme4.* ^51^ Student scores were nested within schools via a random intercept. Hypothesised intervention effects were tested via interactions between treatment and time.

As pre-registered, we conducted sensitivity analyses to assess whether findings were robust when controlling for demographic variables. We also conducted a per-protocol analysis by assessing whether completion of the professional learning moderated the effect of the intervention on the primary outcome. Rather than using an arbitrary cut-off, we used learning analytics to identify the percentage of the course that teachers completed at post-test. When students changed teachers between Year 1 and Year 2, we chose the Year 1 teacher completion rate because those teachers spent more time with that student (12 months versus 6-9 months). We assessed whether this percentage explained variance in the effect of the program on their students.

Two exploratory analyses were added after pre-registration. School-based physical activity interventions have typically worked better for boys than for girls ^52^, including trials of iPLAY among mainstream students.^16^ We therefore added a moderation analysis for gender, across all outcomes. Some intervention components may lend themselves better toward acquisition of different fundamental movement skills, so we also conducted a moderation analysis for the subscale (locomotor skills vs. object control) on the primary outcome.

### Qualitative Interviews with Teachers

We invited all teachers and principals from intervention schools to participate in a brief interview about their experiences of the program. Questions were oriented around the barriers and facilitators they experienced, and the key changes they implemented. We provided staff with three options to participate: face-to-face at their school, Zoom interview with researcher, or asynchronous interview via Phonic.ai. Teachers and principals were offered $50 AUD for a 20-30 minute interview. We conducted three rounds of recruitment for these interviews (as many as ethics approval would permit) across 2021 and 2022. During 2021, schools were impacted by COVID-19 restrictions and 2022 was over two years since teachers started the program. As a result, only two teachers consented, and no principals.

## Results

### Recruitment

Recruitment took place between October 2019 and March 2020. Twenty-five schools consented but 5 withdrew before allocation and baseline data collection because of uncertainty around COVID-19. New South Wales schools were instructed to move toward online learning on 23 March 2020.^53^ One school withdrew after baseline data collection and allocation (see Figure 1). In total, we collected baseline data from 214 students from 20 schools (10 control schools, 10 intervention schools). After 21 months, approximately 33% of participants left participating schools, usually to enter high school.

**Figure 1.**
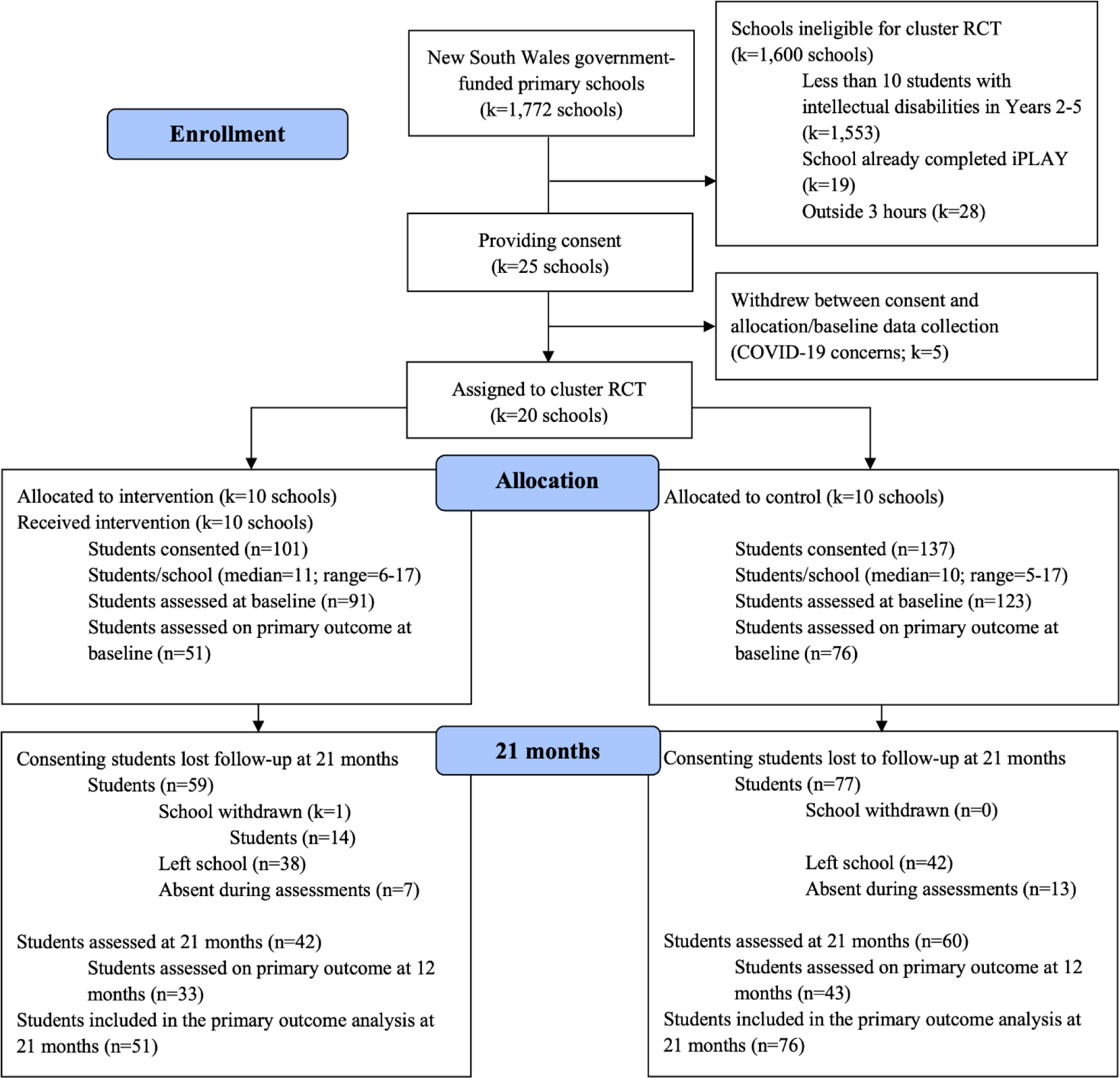
CONSORT flow diagram indicating participant flow throughout the procedure.

Participant characteristics were similar across arms (Table 1), with baseline differences only present for self-concept.

**Table 1.** Characteristics of Control and Intervention Arms at Baseline.

| Characteristic | Control | Intervention | p-value |
| --- | --- | --- | --- |
| Students (n) | 123 | 91 |  |
| Schools (n) | 10 | 10 |  |
| Female (%) | 25.20 | 28.57 | .746 |
| Australian born (%) | 92.68 | 92.31 | .999 |
| English not spoken at home (%) | 10.57 | 7.69 | .607 |
| Functional Movement Skill points | 16.14 (9.53) | 14.78 (10.93) | .471 |
| MVPA (mins) | 73.39 (30.52) | 65.12 (30.46) | .179 |
| MVPA school (mins) | 27.44 (12.90) | 23.19 (12.26) | .082 |
| MVPA school breaks (mins) | 12.00 (7.35) | 12.00 (7.80) | .998 |
| MVPA after school (mins) | 29.94 (15.81) | 24.37 (16.93) | .106 |
| MVPA weekends (mins) | 59.99 (36.28) | 54.68 (30.32) | .382 |
| Self-Concept | 4.23 (0.78) | 3.89 (0.86) | .007** |
| PE Enjoyment | 4.51 (0.89) | 4.29 (0.90) | .092 |
| Life Satisfaction | 4.52 (0.96) | 4.27 (1.14) | .106 |

*Note.* Significance tests were conducted using t-tests for continuous variables, and chi-squared tests for binary variables. \*\* $p < 0.01$

### Primary Outcome

There were no statistically significant group-by-time effects of the intervention on fundamental movement skill competency (b = 1.07 [95% CI -3.70, 5.84], p = .658; see Figure 1). This pattern of results was consistent across the unadjusted model, and one which controlled for participant demographics (b = 1.23 [95% CI -3.55, 6.01], p = .612; see Supplementary Tables).

Per protocol analyses assessed whether intervention effects were explained by the percentage of the course completed by the teachers. Completion rate was bimodal. Forty-five percent of teachers did more than half of the online modules—of those, the median completion was 88% of the online course. Of those who did less than half the modules, the median completion was only 18% of the online course. These completion rates significantly predicted students’ FMS competency (b = 10.40 [95% CI 1.94, 18.87], p = .016), but did not predict *change* in fundamental movement skills over time (b = 3.81 [95% CI -3.16, 10.79], *p* = .28). As shown in Figure 3, students of teachers who completed the program had higher FMS at baseline and post, but did not improve faster over time.

**Figure 2.**
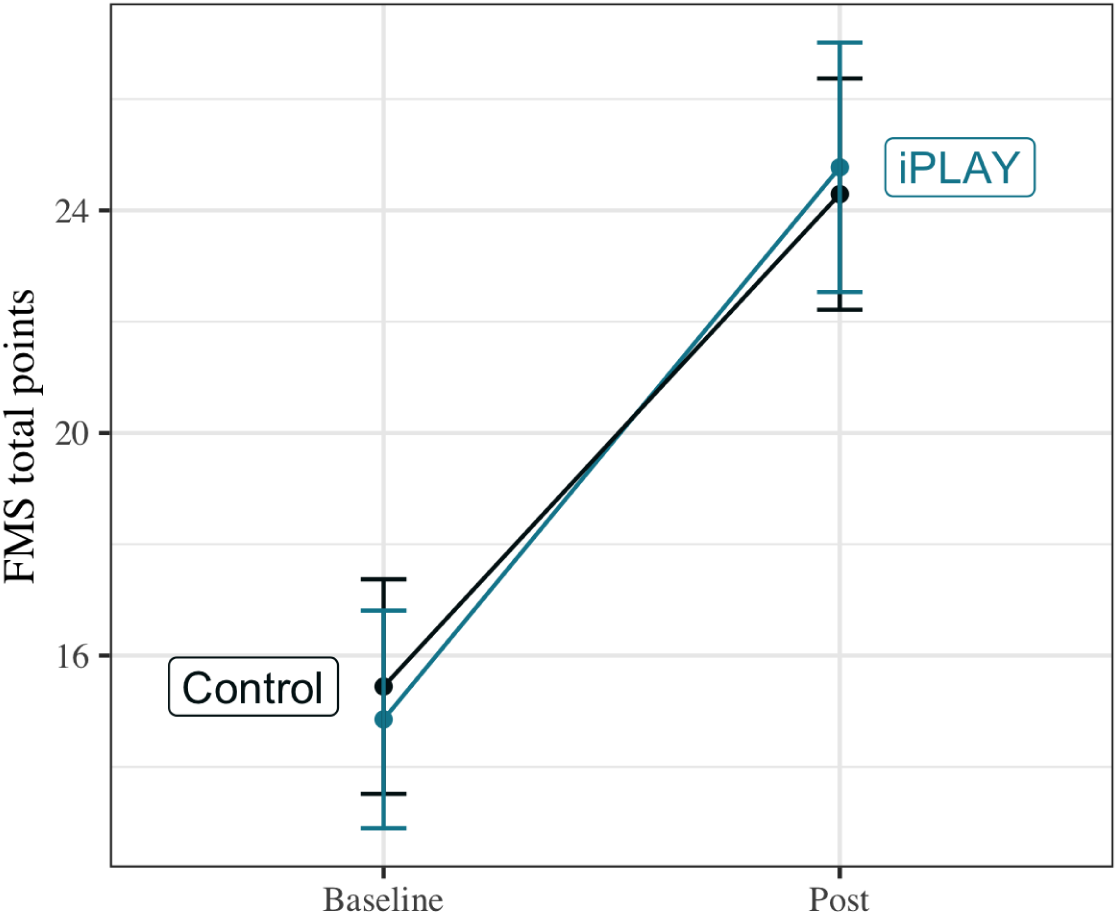
Effect of intervention on fundamental movement skill competency (unadjusted models) *Note*. Error bars show mean ± 1 standard error

**Figure 3.**
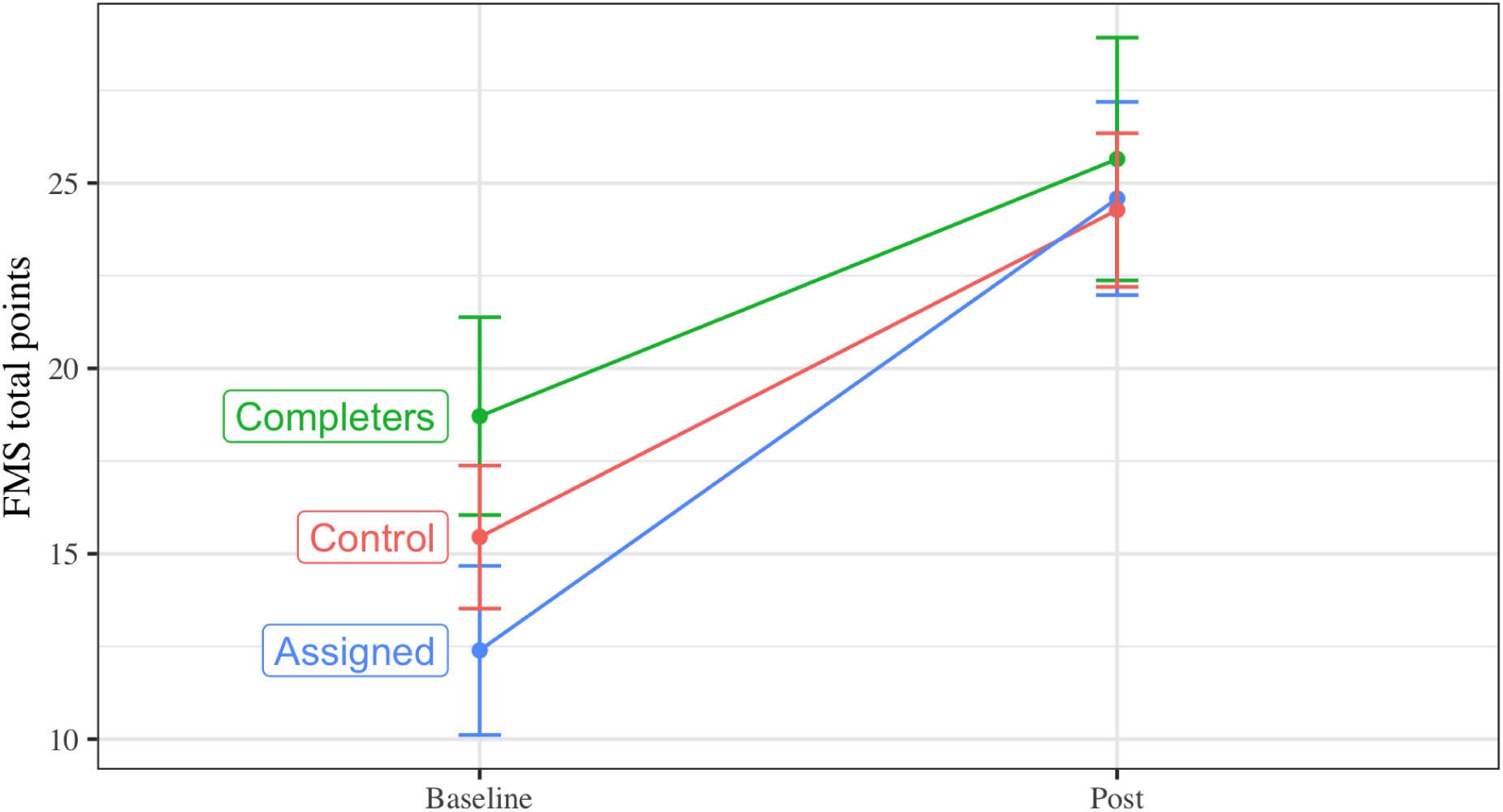
Effect of intervention on fundamental movement skill competency (unadjusted models), with those assigned to the intervention group split by whether or not their teacher completed more than 50% of the program. *Note*. Error bars show mean ± 1 standard error

### Secondary outcomes

As per Table 2, there were no statistically significant effects on the secondary outcomes of cardiorespiratory fitness (via 300-yard run times, b = 0.10 [95% CI -0.36, 0.56], p = .661) student self-concept (b = -0.13 [95% CI -0.62, 0.35], p = .589), PE enjoyment (b = -0.04 [95% CI -0.63, 0.56], p = .90), life satisfaction (b = -0.28 [95% CI -0.93, 0.38], p = .41), or physical activity (e.g., total MVPA b = 8.50 [95% CI -23.83, 40.82], p = .60). Results were consistent when controlling for demographic variables (see supplementary materials).

**Table 2.** Effects of intervention on primary and secondary outcomes.

| *Outcome* | *Intervention effect [95% CI]* | *SE* | *t* | *p* |
| --- | --- | --- | --- | --- |
| Fundamental movement skills | 1.07 [-3.70, 5.83] | 2.42 | 0.44 | .659 |
| Run time | 0.10 [-0.36, 0.56] | 0.23 | 0.44 | .661 |
| MVPA school time | 1.33 [-6.57, 9.23] | 3.97 | 0.34 | .738 |
| MVPA school breaks | -0.65 [-6.01, 4.70] | 2.66 | -0.25 | .808 |
| MVPA after school | 0.76 [-9.41, 10.94] | 5.11 | 0.15 | .882 |
| MVPA weekends | 6.92 [-14.06, 27.89] | 10.53 | 0.66 | .513 |
| MVPA all | 8.50 [-23.83, 40.82] | 16.21 | 0.52 | .602 |
| Self-concept | -0.13 [-0.62, 0.35] | 0.25 | -0.54 | .589 |
| PE enjoyment | -0.04 [-0.63, 0.56] | 0.30 | -0.13 | .900 |
| Life satisfaction | -0.28 [-0.93, 0.38] | 0.33 | -0.83 | .406 |

*Note.* MVPA = Moderate to vigorous physical activity; PE = physical education

### Harms

We had no reports of any students or teachers experiencing harms from their participation in this study.

### Qualitative Interviews with Teachers

Staff found the iPLAY training, mentors, and website helpful (see Supplementary Materials for quotes). They recommended few changes to the content, if any. Major barriers to adoption were competing priorities placed upon teachers (curriculum changes, other professional learning). These competing priorities made it hard to prioritise online, self-paced professional learning, unless supported by their principals. Teachers reported focusing their implementation on classroom energizers and differentiation for physical education lessons (e.g., SAAFE principles and TREE framework).

## Discussion

We hypothesised that a blended-learning teacher professional development program would increase fundamental movement skill competency among primary students with intellectual disability. We did not find the intervention increased children’ fundamental movement skills, or any of our secondary outcomes (enjoyment, wellbeing, self-concept, physical activity, or cardiorespiratory fitness). Completion rates and qualitative data suggest adoption was influenced by the COVID-19 pandemic and competing demands placed on teachers. Effects may also have been reduced by the high quality of teaching present in the comparison group.

Our findings are consistent with previous research on physical activity interventions for children with intellectual disability, where effects are usually small or non-significant.^11^ Programs that are effective in this population appear to have significantly higher costs per participant (e.g., modified bikes; guided resistance training programs).^11^ Among regular students, school-based physical activity interventions are low cost so can be delivered at scale,^16,17^ but they also typically demonstrate modest effects (significant only among some students).^52^ Our study was likely underpowered to detect modest effects.

The data suggest a few other reasons why we may have found no significant benefits. First, around half of teachers who started the program finished the modules. The bimodal distribution may be explained by the incentive structure, where teachers received government-mandated professional learning ‘hours’ for completing the program, but received no hours for doing less. That incentive would explain a bimodal distribution: teachers who needed those hours may have engaged, and others may have already met their quota for hours. It reflects that adoption rates as high as 50% may not generalise to school contexts without these incentives. Alternatively, the bimodal distribution may suggest selection bias in teachers who complete the program. Our data showed teachers who completed the program tended to have students who were more competent at baseline. It is possible that teachers who completed the training are the ones already more engaged in promoting physical activity, or ones teaching students with less severe disability. We do not have data regarding severity of student disability or teacher attitudes toward physical activity, so identifying causes of—and solutions to—this selection bias are important for future research.

Adoption rates like ours are common in the online delivery of professional learning.^54^ Engagement in online professional learning is a known challenge, so we tried to embed many of the recommendations for improving engagement from Lee and colleagues (e.g., emphasising intrinsic value, ensuring user friendliness, asking principals to allocate time).^54^ Nevertheless, our study may reveal a trade off between low-cost interventions with low researcher contact time, and high-touch interventions with higher adoption and effect sizes. Future studies may want to compare the cost-effectiveness of our blended learning model against more intensive models with more synchronous time with teachers, to see if this increases adoption. The adoption rates may also be partially explained by COVID-19, with intervention delivery beginning in a year of lockdowns and remote schooling. Our qualitative data suggested the biggest barriers for teachers were not in the implementation of the iPLAY4i strategies, but that competing demands got in the way of online professional learning. When iPLAY was delivered to regular teachers prior to the pandemic, the same course and incentive structure led to higher completion rates (63% vs 45% here).^16^ So, if this study were replicated, we anticipate adoption to be higher without the interference of a global pandemic.

Effects may have also been modest due to strong improvements even among the control group. As shown in Figure 2, even the control group had a 50% improvement in performance on the TGMD-3, relative to baseline. As shown in supplementary materials, the average student was getting over 60 minutes of moderate-to-vigorous physical activity each school day, meeting WHO guidelines.^5^ These data may represent a ‘Hawthorne effect’, where students, teachers, or parents increased physical activity due to the mere presence of an accelerometer. Alternatively, these data suggest ‘business as usual’ in these schools may be sufficient to help children with disabilities at this age. It appears physical activity most precipitously declines somewhat later in development among this population. For example, Shields et al.^55^ found that differences in activity between young people with and without disabilities become more pronounced in adolescence. Interventions may therefore be more important as students start transitioning into adolescence.^56^ Whole-of-school interventions may also need to target different components to what we included. Our curricular components often focused on adapting lessons to children’s abilities (e.g., via the TREE framework).^30^ This kind of differentiation may be routine for teachers of students with disability; if this kind of adaptation is already routine, training may not be valuable. In contrast, the small amount of qualitative data we had suggested teachers got value from the classroom energizers. It is possible that our intervention could have focused less on improving the quality of physical education, but instead focused on increasing the *opportunity* for students to be active (e.g., via more frequent, brief physical activity sessions).^56^ Future studies should aim to explore current teacher beliefs and knowledge gaps before and after the intervention. Doing so would help assess what teachers are learning most from the professional learning interventions, and what learning best predicts student outcomes.

## Limitations and Future Directions

Our results have a number of important caveats. First, our study was significantly impacted by COVID-19. While it was important for clinical trials to continue despite the pandemic,^25^ government-imposed restrictions meant we needed to delay end-point data collection by three school terms. This delay may have allowed some intervention effects to wash out. It also meant over 30% of our sample left for high school and were lost to follow-up. With this dropout—and a large number of participants who could not, or would not complete our primary outcome—our study was likely underpowered. It also appears our intervention effects were smaller than those from meta-analyses of fundamental movement skill training for typically developing children.^20^ Therefore, while our intervention was low cost and therefore easy to scale, the effects may be smaller than more intensive interventions for students without disability. We also tried to target a range of physical activity outcomes, not only fundamental movement skills. Teachers were encouraged to make physical activity more motivating, more frequent, and better differentiated to student abilities. It is likely that an intervention would have demonstrated stronger effects for our primary outcome if it focused *only* on fundamental movement skills. Future studies may want to compare the outcomes from focused interventions against diffuse interventions like ours, targeting a range of outcomes.

Our primary outcome (fundamental movement skill competency) had blinded, trained researchers using a measure validated among children with intellectual disabilities (TGMD-3).^33–35^ However, many of our other measures may have less good reliability or validity. For example, our self-report measures (e.g., self-concept, subjective wellbeing) were drawn from validated measures, but the response scales might be less sensitive to changes than measures appropriate for typically developing children. This may explain why we found no changes on self-reported measures, but previous evaluations of iPLAY among typically developing students saw gains in wellbeing, motivation, and enjoyment.^17^ Alternatively, children with disability may have a greater number of factors that influence these variables (e.g., direct and indirect consequences of their disability). So, against these background factors, the effects from our professional learning program may be too small to detect. We did not conduct power analyses for these secondary outcomes. Future studies may want to better estimate the influence of physical activity on, say, wellbeing and adequately power their design to detect these effects.

## Conclusions

Our study found this school based intervention did not reliably increase fundamental movement skills, physical activity, or self-reported outcomes among children with intellectual disability. Effective, scalable school-based physical activity interventions for children with disability remain elusive. Given the percentage of the population with disability, and increased risk of chronic disease among this population, researchers should continue exploring novel ways of increasing physical activity and physical literacy among this population.

## Supporting information

CONSORT Checklist

## Protocol Registration

ANZCTR registration number: ACTRN12620000405910

## Data availability statement

Deidentified data and code for reproducing the analyses are available on the Open Science Framework: https://osf.io/jqt32

## Acknowledgements

We express our sincere gratitude to Kirsty Bergan for her role in the project.

## References

1. World Health Organisation. World Report on Disability. World Health Organisation; 2011. Accessed February 7, 2024. https://www.who.int/teams/noncommunicable-diseases/sensory-functions-disability-and-rehabilitation/world-report-on-disability

2. Thompson JR, Shogren K, Wehmeyer M. Supports and support needs in strengths-based models of intellectual disability. Published online November 11, 2016:39–57. doi:10.4324/9781315736198.CH3

3. O’Leary L, Cooper SA, Hughes-McCormack L. Early death and causes of death of people with intellectual disabilities: A systematic review. J Appl Res Intellect Disabil. 2018;31(3):325–342. doi:10.1111/jar.12417

4. World Health Organisation. Global Report on Health Equity for Persons with Disabilities. World Health Organisation; 2022. Accessed February 7, 2024. https://www.who.int/teams/noncommunicable-diseases/sensory-functions-disability-and-rehabilitation/global-report-on-health-equity-for-persons-with-disabilities

5. World Health Organisation. WHO Guidelines on Physical Activity and Sedentary Behaviour. World Health Organisation; 2020. Accessed February 7, 2024. https://www.who.int/publications/i/item/9789240015128

6. Aubert S, Barnes JD, Demchenko I, et al. Global Matrix 4.0 Physical Activity Report Card grades for children and adolescents: Results and analyses from 57 countries. J Phys Act Health. 2022;19(11):700–728. doi:10.1123/jpah.2022-0456

7. Ross SM, Smit E, Yun J, Bogart K, Hatfield B, Logan SW. Updated National Estimates of Disparities in Physical Activity and Sports Participation Experienced by Children and Adolescents With Disabilities: NSCH 2016-2017. J Phys Act Health. 2020;17(4):443–455. doi:10.1123/jpah.2019-0421

8. Martin Ginis KA, van der Ploeg HP, Foster C, et al. Participation of people living with disabilities in physical activity: a global perspective. Lancet. 2021;398(10298):443–455. doi:10.1016/S0140-6736(21)01164-8

9. Kapsal NJ, Dicke T, Morin AJS, et al. Effects of Physical Activity on the Physical and Psychosocial Health of Youth With Intellectual Disabilities: A Systematic Review and Meta-Analysis. J Phys Act Health. 2019;16(12):1187–1195. doi:10.1123/jpah.2018-0675

10. World Health Organisation. Promoting Physical Activity through Schools: A Toolkit. World Health Organisation; 2021. Accessed February 7, 2024. https://www.who.int/publications/i/item/9789240035928

11. McGarty AM, Downs SJ, Melville CA, Harris L. A systematic review and meta-analysis of interventions to increase physical activity in children and adolescents with intellectual disabilities: Effect of interventions on physical activity. J Intellect Disabil Res. 2018;62(4):312–329. doi:10.1111/jir.12467

12. Bloemen M, Van Wely L, Mollema J, Dallmeijer A, de Groot J. Evidence for increasing physical activity in children with physical disabilities: a systematic review. Dev Med Child Neurol. 2017;59(10):1004–1010. doi:10.1111/dmcn.13422

13. Hinckson EA, Dickinson A, Water T, Sands M, Penman L. Physical activity, dietary habits and overall health in overweight and obese children and youth with intellectual disability or autism. Res Dev Disabil. 2013;34(4):1170–1178. doi:10.1016/j.ridd.2012.12.006

14. Sun Y, Yu S, Wang A, et al. Effectiveness of an adapted physical activity intervention on health-related physical fitness in adolescents with intellectual disability: a randomized controlled trial. Sci Rep. 2022;12(1):22583. doi:10.1038/s41598-022-26024-1

15. Cohen KE, Morgan PJ, Plotnikoff RC, Callister R, Lubans DR. Physical activity and skills intervention: SCORES cluster randomized controlled trial. Med Sci Sports Exerc. 2015;47(4):765–774. doi:10.1249/MSS.0000000000000452

16. Lonsdale C, Sanders T, Parker P, et al. Effect of a Scalable School-Based Intervention on Cardiorespiratory Fitness in Children: A Cluster Randomized Clinical Trial. JAMA Pediatr. 2021;175(7):680–688. doi:10.1001/jamapediatrics.2021.0417

17. Lubans DR, Sanders T, Noetel M, et al. Scale-up of the Internet-based Professional Learning to help teachers promote Activity in Youth (iPLAY) intervention: a hybrid type 3 implementation-effectiveness trial. Int J Behav Nutr Phys Act. 2022;19(1):141. doi:10.1186/s12966-022-01371-4

18. Damschroder LJ, Aron DC, Keith RE, Kirsh SR, Alexander JA, Lowery JC. Fostering implementation of health services research findings into practice: a consolidated framework for advancing implementation science. Implement Sci. 2009;4(1):50. doi:10.1186/1748-5908-4-50

19. Australian Curriculum, Assessment and Reporting Authority. School students with disability. Australian Curriculum, Assessment and Reporting Authority. Accessed February 9, 2024. https://www.acara.edu.au/reporting/national-report-on-schooling-in-australia/school-students-with-disability

20. Morgan PJ, Barnett LM, Cliff DP, et al. Fundamental movement skill interventions in youth: a systematic review and meta-analysis. Pediatrics. 2013;132(5):e1361–e1383. doi:10.1542/peds.2013-1167

21. Cornish K, Fox G, Fyfe T, Koopmans E, Pousette A, Pelletier CA. Understanding physical literacy in the context of health: a rapid scoping review. BMC Public Health. 2020;20(1):1–19. doi:10.1186/s12889-020-09583-8

22. Maïano C, Hue O, April J. Fundamental movement skills in children and adolescents with intellectual disabilities: A systematic review. J Appl Res Intellect Disabil. Published online May 13, 2019. doi:10.1111/jar.12606

23. Lubans DR, Morgan PJ, Cliff DP, Barnett LM, Okely AD. Fundamental Movement Skills in Children and Adolescents. Sports Med. 2010;40(12):1019–1035. doi:10.2165/11536850-000000000-00000

24. Holfelder B, Schott N. Relationship of fundamental movement skills and physical activity in children and adolescents: A systematic review. Psychol Sport Exerc. 2014;15(4):382–391. doi:10.1016/j.psychsport.2014.03.005

25. McDermott MM, Newman AB. Preserving Clinical Trial Integrity During the Coronavirus Pandemic. JAMA. 2020;323(21):2135. doi:10.1001/jama.2020.4689

26. Noetel M, Lonsdale C. Internet-Based Professional Learning to Help Teachers Promote Activity in Youth with Intellectual Disability. The iPLAY For Inclusion Project: iPLAY4i. Australian New Zealand Clinical Trials Registry. 2020. https://www.anzctr.org.au/Trial/Registration/TrialReview.aspx?id=379417&isReview=tru e

27. Lonsdale C, Sanders T, Cohen KE, et al. Scaling-up an efficacious school-based physical activity intervention: Study protocol for the “Internet-based Professional Learning to help teachers support Activity in Youth” (iPLAY) cluster randomized controlled trial and scale-up implementation evaluation. BMC Public Health. 2016;16(1):873. doi:10.1186/s12889-016-3243-2

28. MacDonald A, McGill P. Outcomes of Staff Training in Positive Behaviour Support: A Systematic Review. J Dev Phys Disabil. 2013;25(1):17–33. doi:10.1007/s10882-012-9327-8

29. Ok MW, Rao K, Bryant BR, McDougall D. Universal Design for Learning in Pre-K to Grade 12 Classrooms: A Systematic Review of Research. Exceptionality. 2017;25(2):116–138. doi:10.1080/09362835.2016.1196450

30. Australian Sports Commission. Using TREE. Sports Ability. May 26, 2019. Accessed May 28, 2024. https://www.sportaus.gov.au/sports_ability/using_tree

31. Lubans DR, Lonsdale C, Cohen K, et al. Framework for the design and delivery of organized physical activity sessions for children and adolescents: rationale and description of the ‘SAAFE’ teaching principles. Int J Behav Nutr Phys Act. 2017;14(1):24. doi:10.1186/s12966-017-0479-x

32. Webster EK, Ulrich DA. Evaluation of the psychometric properties of the Test of Gross Motor Development—third edition. Journal of Motor Learning and Development. 2017;5(1):45–58. https://journals.humankinetics.com/view/journals/jmld/5/1/article-p45.xml

33. Allen KA, Bredero B, Van Damme T, Ulrich DA, Simons J. Test of Gross Motor Development-3 (TGMD-3) with the Use of Visual Supports for Children with Autism Spectrum Disorder: Validity and Reliability. J Autism Dev Disord. 2017;47(3):813–833. doi:10.1007/s10803-016-3005-0

34. Simons J, Eyitayo G. Aspects of reliability and validity of the TGMD-3 in 7-10 year old children with intellectual disability in Belgium. European Psychomotricity Journal. 2016;8(1):3–16. https://lirias.kuleuven.be/1689672?limo=0

35. Magistro D, Piumatti G, Carlevaro F, et al. Measurement invariance of TGMD-3 in children with and without mental and behavioral disorders. Psychol Assess. 2018;30(11):1421–1429. doi:10.1037/pas0000587

36. Simons J, Daly D, Theodorou F, Caron C, Simons J, Andoniadou E. Validity and reliability of the TGMD-2 in 7-10-year-old Flemish children with intellectual disability. Adapt Phys Activ Q. 2008;25(1):71–82. doi:10.1123/apaq.25.1.71

37. Oppewal A, Hilgenkamp TIM, van Wijck R, Evenhuis HM. Cardiorespiratory fitness in individuals with intellectual disabilities--a review. Res Dev Disabil. 2013;34(10):3301–3316. doi:10.1016/j.ridd.2013.07.005

38. Baumgartner T, Horvat M. Reliability of Field Based Cardiovascular Fitness Running Tests for Individuals with Mental Retardation. Adapt Phys Activ Q. 1991;8(2):107–114. doi:10.1123/apaq.8.2.107

39. Aufsesser PM. Effects of repeated testing on the reliability of fitness scores of institutionalized mentally retarded individuals. Am J Ment Defic. 1979;84(3):313–315. https://www.ncbi.nlm.nih.gov/pubmed/525662

40. Leung W, Siebert EA, Yun J. Measuring physical activity with accelerometers for individuals with intellectual disability: A systematic review. Res Dev Disabil. 2017;67:60–70. doi:10.1016/j.ridd.2017.06.001

41. Motl RW, Dishman RK, Saunders R, Dowda M, Felton G, Pate RR. Measuring enjoyment of physical activity in adolescent girls. Am J Prev Med. 2001;21(2):110–117. doi:10.1016/s0749-3797(01)00326-9

42. Tracey DK, Marsh HW. Self-concepts of primary students with mild intellectual disabilities: Issues of measurement and educational placement. Self-concept theory, research and practice: Advances for the new millennium. Published online 2000:419. http://citeseerx.ist.psu.edu/viewdoc/download?doi=10.1.1.195.6552&rep=rep1&type=pdf#page=427

43. Marsh HW, Ellis LA, Parada RH, Richards G, Heubeck BG. A short version of the Self Description Questionnaire II: operationalizing criteria for short-form evaluation with new applications of confirmatory factor analyses. Psychol Assess. 2005;17(1):81–102. doi:10.1037/1040-3590.17.1.81

44. Cheung F, Lucas RE. Assessing the validity of single-item life satisfaction measures: results from three large samples. Qual Life Res. 2014;23(10):2809–2818. doi:10.1007/s11136-014-0726-4

45. Faul F, Erdfelder E, Lang AG, Buchner A. G*Power 3: a flexible statistical power analysis program for the social, behavioral, and biomedical sciences. Behav Res Methods. 2007;39(2):175–191. https://www.ncbi.nlm.nih.gov/pubmed/17695343

46. Bland JM. Cluster randomised trials in the medical literature: two bibliometric surveys. BMC Med Res Methodol. 2004;4:21. doi:10.1186/1471-2288-4-21

47. Moore RT, Schnakenberg K. Block, Assign, and Diagnose Potential Interference in Randomized Experiments.; 2016. https://cran.r-project.org/web/packages/blockTools/blockTools.pdf

48. R Core Team. R: A Language and Environment for Statistical Computing. R Foundation for Statistical Computing; 2022. https://www.R-project.org/

49. Twisk JW, Rijnhart JJ, Hoekstra T, Schuster NA, Ter Wee MM, Heymans MW. Intention-to-treat analysis when only a baseline value is available. Contemp Clin Trials Commun. 2020;20(100684):100684. doi:10.1016/j.conctc.2020.100684

50. van Buuren S, Groothuis-Oudshoorn K. mice: Multivariate Imputation by Chained Equations in R. J Stat Softw. 2011;45(3):1–67. doi:10.18637/jss.v045.i03

51. Bates D, Mächler M, Bolker B, Walker S. Fitting Linear Mixed-Effects Models Using lme4. *Journal of Statistical Software*, Articles. 2015;67(1):1–48. doi:10.18637/jss.v067.i01

52. Hartwig TB, Sanders T, Vasconcellos D, et al. School-based interventions modestly increase physical activity and cardiorespiratory fitness but are least effective for youth who need them most: an individual participant pooled analysis of 20 controlled trials. Br J Sports Med. Published online January 13, 2021. doi:10.1136/bjsports-2020-102740

53. Clark S. COVID-19: chronology of state and territory announcements on schools and early childhood education in 2020. Parliment of Australia. 2022. Accessed May 8, 2024. https://www.aph.gov.au/About_Parliament/Parliamentary_Departments/Parliamentary_Library/pubs/rp/rp2122/Chronologies/COVID-19-StateTerritoryAnnouncementsSchoolsEducation

54. Lee J, Sanders T, Antczak D, et al. Influences on user engagement in online professional learning: A narrative synthesis and meta-analysis. Rev Educ Res. Published online March 9, 2021:0034654321997918. doi:10.3102/0034654321997918

55. Shields N, Synnot AJ, Barr M. Perceived barriers and facilitators to physical activity for children with disability: a systematic review. Br J Sports Med. 2012;46(14):989–997. doi:10.1136/bjsports-2011-090236

56. Kable TJ, Leahy AA, Smith JJ, et al. Time-efficient physical activity intervention for older adolescents with disability: rationale and study protocol for the Burn 2 Learn adapted (B2La) cluster randomised controlled trial. BMJ Open. 2022;12(8):e065321. doi:10.1136/bmjopen-2022-065321

